# Cross-Device Field-of-View Scale Differences in Clinical OCT Angiography: Phantom Calibration and Large-Scale Clinical Validation

**DOI:** 10.64898/2026.09.04.26361850

**Authors:** Yi Zhang, Zhaoyu Gong, Viet-Hoan Le, Bhagavath Sivathanu Kumar, Yaping Shi, Siyuan Wang, Yu Jiang, Christina Duong, Sharon Henry, Crystal S. Yun, Yue Wu, Aaron Y. Lee, Cecilia S. Lee, Ruikang K. Wang

## Abstract

**Purpose:** Device-specific field-of-view (FOV) may affect quantitative optical coherence tomography angiography (OCTA) metrics. This study quantifies inter-device FOV scale differences across four commonly used clinical OCTA devices.

**Methods:** Four devices were evaluated under a nominal 6×6 mm (or 20°×20°) scanning protocol: Zeiss Cirrus, Heidelberg Spectralis, Topcon Triton, and Topcon Maestro2. Device-specific spatial scale was assessed by (1) phantom calibration using a model eye with a 1×1 mm checkerboard, computing scale factors relative to Zeiss Cirrus; and (2) in vivo validation, registering same-eye, same-visit retinal images from the three devices to those of Zeiss Cirrus and extracting scale factors from the transformation matrices. Correlation between DICOM-implied FOV and spherical equivalent (SE) was assessed, where DICOM-implied FOV equals pixel-spacing × image-dimension.

**Results:** Phantom measurements showed that Spectralis, Triton, and Maestro2 each captured a systematically smaller FOV than Cirrus, with geometric mean scale factors of 0.933, 0.954, and 0.967. The same pattern was observed across 801 eyes from 556 participants, giving geometric mean scale factors of 0.939, 0.943, and 0.960, agreeing with phantom estimates within 1.2%. DICOM-implied FOV was fixed at 6.000×6.000 mm for three devices but varied from 5.212×5.209 to 6.743×6.739 mm for Spectralis, correlating with SE (ρ = −0.635, P < 0.001).

**Conclusion:** Systematic FOV discrepancies exist across clinical OCTA devices despite nominally identical protocols, and DICOM pixel-spacing should be interpreted with caution as a spatial measure.

**Translational Relevance:** The reported device-specific scale factors could serve as magnification corrections for quantitative OCTA metrics, addressing the systematic spatial inter-device discrepancies.

## Introduction

Optical coherence tomography (OCT) and OCT angiography (OCTA) enable non-invasive, three-dimensional, depth-resolved visualization of retinal and choroidal structure and blood flow without the use of contrasting agents.^1–3^ OCTA has been widely adopted in ophthalmology for the diagnosis, monitoring, and treatment planning of various diseases, including age-related macular degeneration (AMD), diabetic retinopathy, glaucoma, and retinal vein occlusion, through quantitative OCTA metrics, such as foveal avascular zone (FAZ) area, non-perfusion area, vascular parameters, and geographic atrophy (GA) lesion size.^4–8^

The use of multiple OCTA devices is common in routine clinical practice and research.^9–11^ Participants may be imaged on different platforms over time within a single institution,^9,11^ and multicenter studies or referral networks frequently aggregate data from different devices.^11,12^ Furthermore, an increasing number of published OCTA studies pool or directly compare data across devices.^9,10,13^ However, inter-device discrepancies in quantitative OCTA metrics are well documented ^10,14^ and could limit direct comparison.

These discrepancies have been mainly attributed to differences in signal processing, including angiographic contrast methods and slab segmentation definitions.^10,15,16^ Accordingly, standardization efforts have mostly focused on algorithmic harmonization.^17–19^ However, an underappreciated source is spatial scaling inconsistency across devices. In particular, the actual field of view (FOV) can deviate from the nominal FOV. The actual FOV can be modeled by the widely used Littmann– Bennett formula,^20^ and it is determined by both a participant-specific factor related to axial length and a device-specific scale factor related to the angular FOV. Variation in either factor, across participants or across devices, can therefore produce magnification-related discrepancies between the actual and nominal FOV that propagate directly into length-or area-based quantitative measurements.^9,21,22^ Although magnification correction has been suggested as one of the key priorities in OCTA standardization,^9^ only 8% of published studies apply it before quantitative analysis,^9,23^ and uncorrected magnification error can bias quantitative metrics substantially, for example, introducing errors of up to 51% in FAZ area.^21,24^

Moreover, among those few studies that apply magnification correction, most rely solely on participant axial length adjustments.^21,24–26^ Within a single device, the device-specific scale factor is typically constant because the angular FOV is set by hardware; therefore, axial length correction alone is sufficient. However, in multi-device imaging, device-specific scale differences remain even after axial length correction, making axial length alone insufficient for magnification correction. Despite this importance, whether device-specific scale discrepancies exist across commercially available clinical OCTA platforms under their respective scan settings, particularly in a large cohort, remains to be addressed.

In this study, we systematically evaluate the device-specific spatial scale across four clinical OCTA devices under a common nominal scanning protocol (6×6 mm or 20° × 20°, one of the most widely used scan settings in clinical OCTA practice and research ^27^), namely Zeiss Cirrus HD-OCT 5000, Heidelberg Spectralis, Topcon DRI-OCT Triton, and Topcon Maestro2, using two complementary approaches: (1) phantom grid calibration using a physical model eye with a 1×1 mm checkerboard pattern, which isolates the device-specific scale by removing patient variability; (2) large-scale in vivo validation using same-eye, same-visit cross-device imaging combined with image registration, which captures the scale offset under real-world clinical acquisition conditions. We further examine whether DICOM pixel-spacing metadata, an increasingly common representation of spatial scale in automated OCTA analysis pipelines, accurately reflects the measured device-specific scale, given its potential to propagate device-dependent biases into downstream quantitative measurements.

## Methods

### 2.1 Devices and Scan Protocol

Four commercially available clinical OCTA devices were evaluated: (1) Zeiss Cirrus HD-OCT 5000 with AngioPlex (Carl Zeiss Meditec, Dublin, CA); (2) Heidelberg Spectralis OCT2 with OCT Angiography Module (Heidelberg Engineering, Heidelberg, Germany); (3) Topcon DRI-OCT Triton (Topcon Corporation, Tokyo, Japan); (4) Topcon Maestro2 (Topcon Corporation, Tokyo, Japan). All devices were configured to acquire the standard macular scan protocol using manufacturer-default settings. While Cirrus, Triton, and Maestro2 specify this scan field directly as 6×6 mm, Spectralis defines the equivalent protocol in angular terms as 20°×20°, which is commonly considered equivalent to 6×6 mm.^28^ To enable consistent axis-specific comparison across devices, all images were oriented such that the horizontal axis (X, image width) corresponded to the same nasal–temporal retinal direction and the vertical axis (Y, image height) to the superior–inferior direction. While Cirrus, Triton, and Maestro2 export the images in this orientation natively, Spectralis exports in a rotated orientation. Thus, Spectralis images were rotated to match those from the other three devices in this study.

### 2.2 Phantom Calibration

A Zeiss model eye phantom (Zeiss, Dublin Innovation Center) was used to compare device-specific spatial scale across devices. The phantom consists of an N-BK7 plano-convex optical lens with an effective focal length of 17.17 mm at 750 nm, refractive index 1.5168 ± 0.0005, center thickness 25.20 ± 0.03 mm, and a chrome checkerboard pattern on the rear lens surface consisting of 1×1 mm squares within a 12×12 mm total pattern area (dimensional accuracy ±30 μm). Dimensions are specified in the projected image plane after accounting for the phantom lens.

The phantom was mounted and secured using the same articulating arm for all devices. It was positioned to approximate the location of a patient’s eye during clinical imaging relative to each device’s headrest and scanned per device using the standard 6×6 mm or 20°×20° protocol. As in clinical scanning, focus was adjusted manually by trained operators for the Cirrus, Spectralis, and Triton systems; for Maestro2, focus was set automatically by the device. For each device, the FOV along each axis was measured in 7 separate sessions in ImageJ (NIH, Bethesda, MD) using the checkerboard grid as a physical reference scale. The coefficient of variation (CV = standard deviation/mean × 100%) across the 10 measurements was calculated for measurement repeatability.

Relative linear scale factors were computed for each axis as the ratio of each device’s measured FOV to that of Cirrus. The area scale factor was defined as the product of the X and Y linear scale factors, and the geometric mean scale factor as the square root of the product of the X and Y scale factors, equivalent to the square root of the area scale factor. Cirrus was selected as the reference because it captured the largest FOV during visual inspection of phantom images, providing a natural upper bound. The choice of reference does not affect the conclusion.

### 2.3 AI-READI Clinical Dataset

To further validate device-specific spatial scale in a large clinical dataset, we utilized publicly available Artificial Intelligence Ready and Exploratory Atlas for Diabetes Insights (AI-READI) dataset (v 3.0.0), a multimodal dataset collected across the University of Alabama at Birmingham, the University of California San Diego, and the University of Washington, comprising participants with varying stages of type 2 diabetes mellitus (T2DM).^29,30^ Each participant was imaged on all four devices at the same visit using manufacturer-default acquisition protocols; only the 6×6 mm (20°×20° for Spectralis) macular OCTA protocol was analyzed. En face OCTA images were exported directly from each device. Age was available in the public release; sex was not included due to participant privacy protections. Autorefraction data (sphere and cylinder) were obtained, and the spherical equivalent (SE) was computed as sphere + cylinder/2. Axial length was not directly measured in AI-READI and was estimated from SE using the formula AL = 23.45 − 0.39 × SE. ^31^ This estimate is used here only as an approximation for better interpretation.

#### 2.3.1 Cross-Device Image Registration

Pairwise image registration was performed between Cirrus (reference) and each of the three comparison devices, Spectralis, Triton, and Maestro2, independently. Cirrus was again chosen as the reference for consistency with phantom study. Two graders (Y. Z and S. W) first reviewed each image pair and excluded eyes with poor image quality, such as motion artifacts, poor signal, or disconnected vessel networks in either device. Feature-based registration was then performed on the remaining eyes using KAZE feature extraction ^32–34^, followed by affine transformation estimation with Random Sample Consensus (RANSAC) outlier rejection.^35^ Registration quality was then evaluated through visual inspection of large-vessel alignment between the registered image pairs. A normalized cross-correlation (NCC) score below 0.25 was used as a supplementary quantitative reference to support decision-making in uncertain cases. Image pairs with poor spatial alignment were excluded. Inter-grader agreement was quantified using quadratic weighted Cohen’s kappa, and disagreements were resolved by consensus. Only eyes passing both image and registration quality checks for all three device pairs were retained in the final dataset to ensure all cross-device comparisons were performed on an identical set of eyes.

For each registered pair, the affine transformation matrix was decomposed into a 2×2 linear component 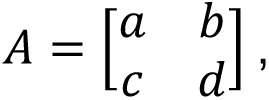 which encodes the relative spatial scaling between the comparison device and Zeiss Cirrus. Axis-specific linear scale factors were calculated as the Euclidean norms of the columns of matrix A. Specifically, the scale factors along the horizontal (*S_X_*) and vertical (*S_Y_*) axes were computed as *S_X_* = 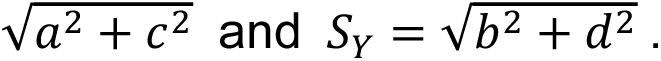 The geometric mean scale factor was defined as 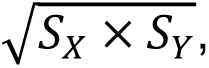 and the area scale factor as *S_X_* × *S_Y_*, following the same definitions as in the phantom calibration. A determinant-based scale factor was additionally obtained by 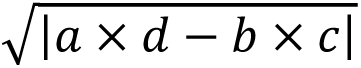 as a rotation-invariant consistency check. Agreement between phantom-and registration-derived scale factors was quantified as the relative percentage deviation of the registration-derived value from the phantom value, computed separately for X, Y, and the geometric mean. Scale factor magnitudes were expressed as percentage deviations using a common convention: linear scale deviation (%) = (Scale_A − Scale_B) / Scale_B × 100, and area scale deviation (%) = (Area Scale_A − Area Scale_B) / Area Scale_B × 100. For comparisons with Cirrus, Scale_B was defined as the scale factor of Cirrus, and Scale_A as that of each non-Cirrus device. For pairwise comparisons among non-Cirrus devices, Scale_B was defined as the scale factor of the device with the larger FOV, and Scale_A as that of the device with the smaller FOV. Inter-subject consistency of registration-derived scale factors was assessed using the coefficient of variation (CV) computed across eyes for each device pair, separately for X, Y, and the geometric mean scale factor.

#### 2.3.2 DICOM Metadata Analysis

We additionally evaluated whether DICOM pixel spacing accurately reflects the device-specific spatial scale observed in AI-READI. DICOM pixel-spacing tags were extracted per eye per device. DICOM-implied FOV was computed as the product of the stored image dimension and the corresponding pixel spacing value, calculated independently for X and Y. The relationship between DICOM-implied FOV and SE or estimated axial length was additionally examined.

### 2.4 Statistical Analysis

All statistical analyses were performed in Python (v3.11.7). Continuous variables are reported as mean ± SD. Given the large sample size (N = 801), the sampling distributions of the test statistics are approximately normal by the central limit theorem; therefore, formal normality tests were not performed. Statistical significance was defined as P < 0.01.

Registration-derived geometric mean scale factors were tested against the null value of 1.0 using one-sample t-tests. Axis-specific scale factors along the X and Y axes were tested against 1.0 using one-sample t-tests with Bonferroni correction across six tests (three devices × two axes). Differences in geometric mean scale factors across the three comparison devices were assessed globally using one-way ANOVA, followed by pairwise two-sample t-tests with Bonferroni correction across three comparisons. Sensitivity analyses were performed to assess whether age and diabetes status confounded the registration-derived scale factors. Linear mixed-effects models were fitted with geometric mean scale factors as the dependent variable, age and diabetes status as fixed effects, and a random intercept per patient to account for within-patient correlation between bilateral eyes. Spearman’s rank correlation coefficients were computed between DICOM-implied FOV and SE, and between DICOM-implied FOV and estimated axial length, within each device given the wide refractive range in the subjects.

## Results

### 3.1 Phantom Calibration

Figure 1 shows representative en face OCTA images of the eye phantom acquired by the four devices, with visible FOV differences across devices. Phantom-measured FOV is summarized in Table 1.

**Figure 1.**
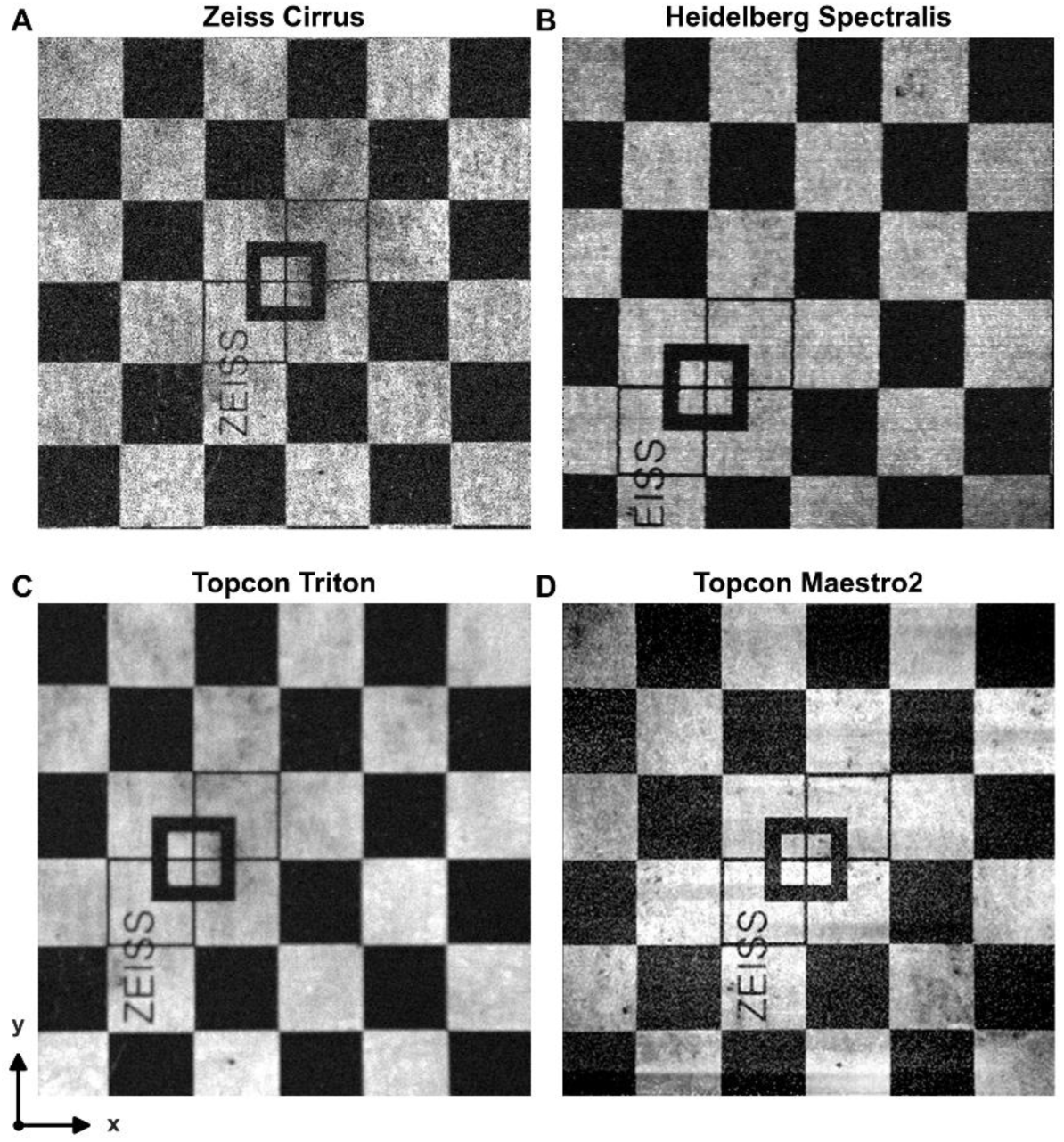
Representative en face OCTA images of the Zeiss model eye phantom acquired by each device under the standard 6×6 mm (or 20°×20°) protocol: (A) Zeiss Cirrus, (B) Heidelberg Spectralis, (C) Topcon Triton, (D) Topcon Maestro2. Axes indicate horizontal (X) and vertical (Y) directions. The images are directly exported from the machine with only brightness adjustment.

**Table 1.** Phantom-Measured Field of View and Spatial Scale Factors Across OCTA Devices.

| Device | FOV X,<br>mm<br>(Mean ±<br>SD) | CV<br>X, % | FOV Y,<br>mm<br>(Mean ±<br>SD) | CV<br>Y, % | Scale<br>X | Scale<br>Y | Geometric<br>Mean<br>Scale | Area<br>Scale |
| --- | --- | --- | --- | --- | --- | --- | --- | --- |
| Zeiss Cirrus<br>(reference) | 6.012 ±<br>0.023 | 0.38 | 6.076 ±<br>0.014 | 0.23 | — | — | — | — |
| Heidelberg<br>Spectralis | 5.690 ±<br>0.010 | 0.18 | 5.598 ±<br>0.010 | 0.18 | 0.946 | 0.921 | 0.933 | 0.871 |
| Topcon Triton | 5.787 ±<br>0.012 | 0.21 | 5.733 ±<br>0.016 | 0.28 | 0.963 | 0.944 | 0.954 | 0.909 |
| Topcon<br>Maestro2 | 5.881 ±<br>0.010 | 0.17 | 5.808 ±<br>0.011 | 0.19 | 0.978 | 0.956 | 0.967 | 0.935 |

Scale X and Scale Y is relative to Zeiss Cirrus; Geometric mean scale = $\sqrt{(\text{Scale X} \times \text{Scale Y})}$ ; Area Scale = Scale X × Scale Y.
FOV was measured 7 times per device along each axis. CV = $(\text{SD}/\text{mean}) \times 100\%$ , reflecting measurement reproducibility. CV, coefficient of variation; FOV, field of view; SD, standard deviation.

Using the current eye phantom (Table 1), Cirrus yielded a measured FOV of 6.012 ± 0.023 mm (X) and 6.076 ± 0.014 mm (Y). The remaining three devices exhibited systematically smaller FOVs. Heidelberg Spectralis showed the largest difference, measuring 5.690 ± 0.010 mm (X) and 5.598 ± 0.010 mm (Y), corresponding to device-specific linear scale factors of 0.946 (X) and 0.921 (Y), a geometric mean scale of 0.933, and an area scale of 0.871, indicating a 12.90% reduction in captured imaging area relative to Cirrus. Triton measured 5.787 ± 0.012 mm (X) by 5.733 ± 0.016 mm (Y), yielding linear scale factors of 0.963 and 0.944, a geometric mean scale of 0.954, and an area scale of 0.909 (9.10% area reduction). Similarly, Maestro2 measured 5.881 ± 0.010 mm (X) by 5.808 ± 0.011 mm (Y), corresponding to linear scale factors of 0.978 (X) and 0.956 (Y), a geometric mean scale of 0.967, and an area scale of 0.935 (6.50% area reduction) relative to Cirrus. Measurement reproducibility was excellent, with CVs below 0.4% across all devices and axes. While absolute FOVs depend on the eye phantom’s axial length, the relative scale factors reported in Table 1 are independent of phantom optical geometry and are therefore directly applicable to clinical imaging.

### 3.2 AI-READI Clinical Dataset

Figure 2 shows representative same-eye OCTA images acquired across the four devices in the AI-READI dataset. Despite the nominally identical 6×6 mm or 20°×20° scanning protocol, the captured retinal areas differ visibly across devices relative to Zeiss.

**Figure 2.**
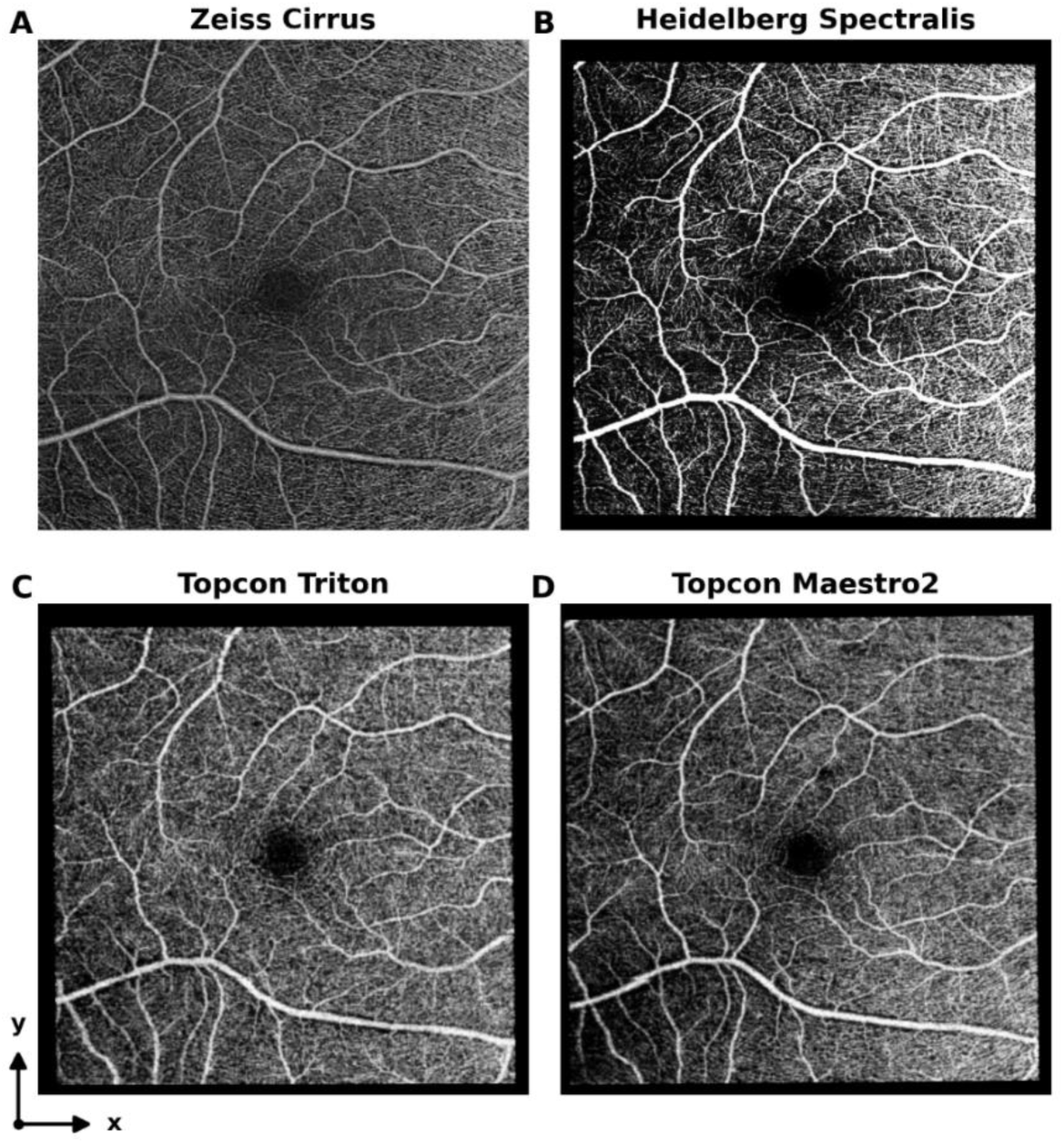
Representative same-eye OCTA en face images across the four devices from an AI-READI participant in their 60s: (A) Zeiss Cirrus, (B) Heidelberg Spectralis, (C) Topcon Triton, (D) Topcon Maestro2. Images represent the superficial retinal slab as exported directly from each device’s proprietary software. Despite nominally identical 6×6 mm (or 20°×20°) protocols, captured retinal areas differ visibly relative to Zeiss. Black borders in (B–D) reflect image padding.

#### 3.2.1 Sample and Demographics

A total of 2197 pairs were identified for Spectralis, 4082 for Triton, and 4143 for Maestro2, each paired against Cirrus. Inter-grader agreement was excellent across all device pairs, with 95% CIs of weighted κ ranging from 0.90 to 0.93. Pairs were excluded for poor image quality, removing 384, 1603, and 1934 pairs for the three comparison devices, respectively, followed by a further 160, 432, and 408 pairs excluded for registration failure or poor alignment. This yielded 1653, 2047, and 1801 pairs passing all criteria. Eyes passing both quality checks across all three device pairs were retained for the final cohort, resulting in 801 eyes from 556 participants. Mean patient age was 61.1 ± 10.7 years. Autorefraction data were available for 274 eyes from 201 patients; within this subset, the SE was −0.60 ± 1.97 D, and the estimated axial length was 23.68 ± 0.77 mm.

#### 3.2.2 Cross-Device Image Registration

Registration-derived geometric mean scale factors were significantly less than 1.0 for all three comparison devices, with all P values below 0.001, indicating a systematically smaller FOV relative to Zeiss Cirrus. As shown in Figure 3, Maestro2 showed the smallest difference at 0.960 (−4.00% linear deviation), followed by Triton at 0.943 (−5.70% linear deviation) and Spectralis at 0.939 (−6.10% linear deviation). The narrow geometric mean scale factor distributions across eyes, with CV ≤ 1.02%, confirmed that the spatial scale offset is a stable device-level property rather than patient-dependent variation. The determinant-based scale factor differed from the geometric mean by less than 0.001% across all devices, indicating that rotation did not meaningfully affect scale factor estimation. Registration-derived values agreed with phantom-measured values within 1.2%, differing by +0.64% for Spectralis, −1.15% for Triton, and −0.72% for Maestro2. Detailed scale factors are provided in Table 2a.

**Figure 3.**
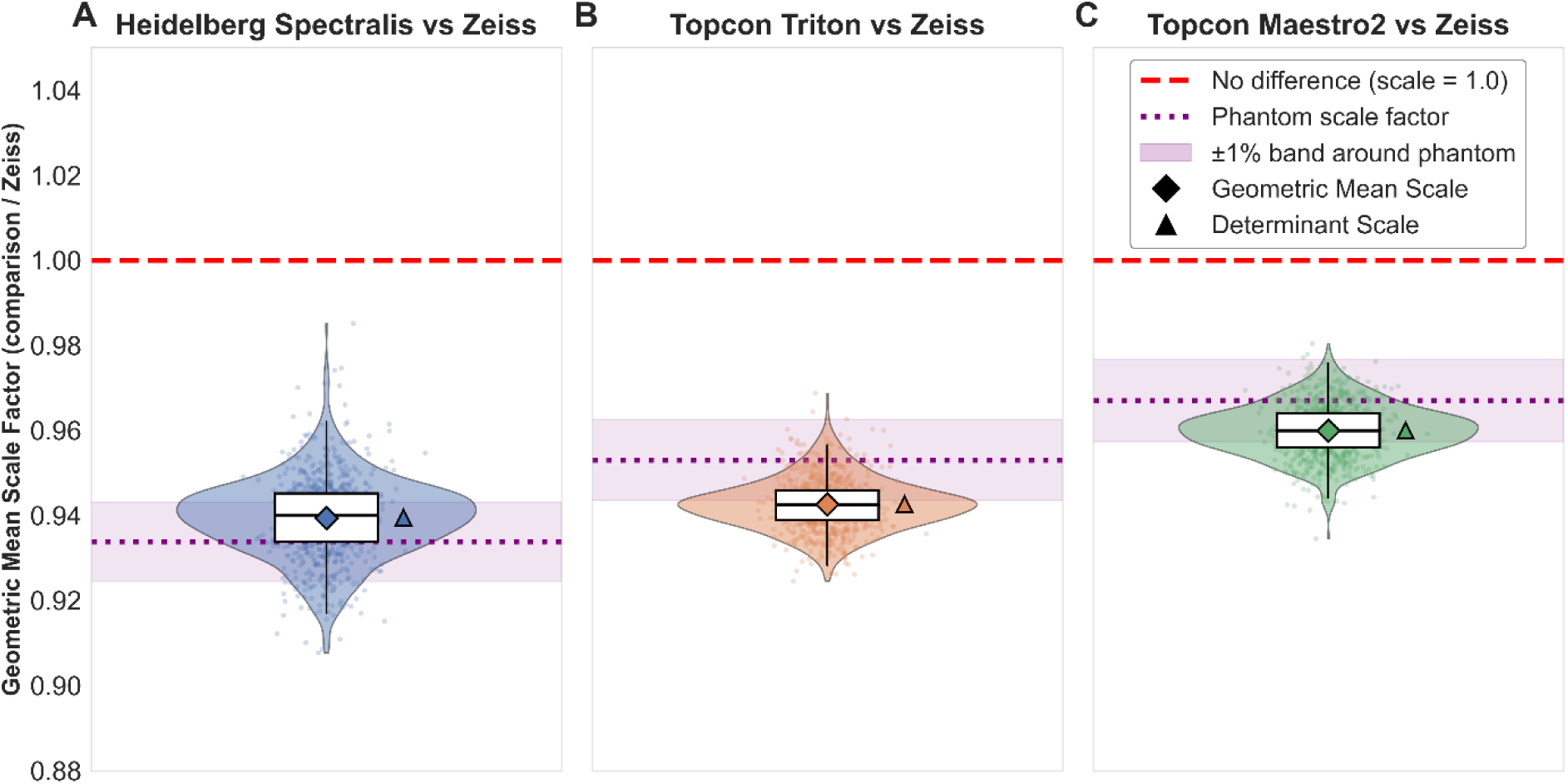
Distribution of registration-derived geometric mean scale factors across 801 eyes for each device pair relative to Zeiss Cirrus: (A) Heidelberg Spectralis, (B) Topcon Triton, (C) Topcon Maestro2. Boxes indicate interquartile range with median, and whiskers are 1.5× IQR. Diamonds mark the geometric mean scale factor; triangles mark the determinant-based equivalent. The red dashed line marks scale = 1.0; the purple dotted line marks the phantom-derived scale factor with a ±1% shaded band. Statistical details in Tables 2a and 2b.

**Table 2a.**
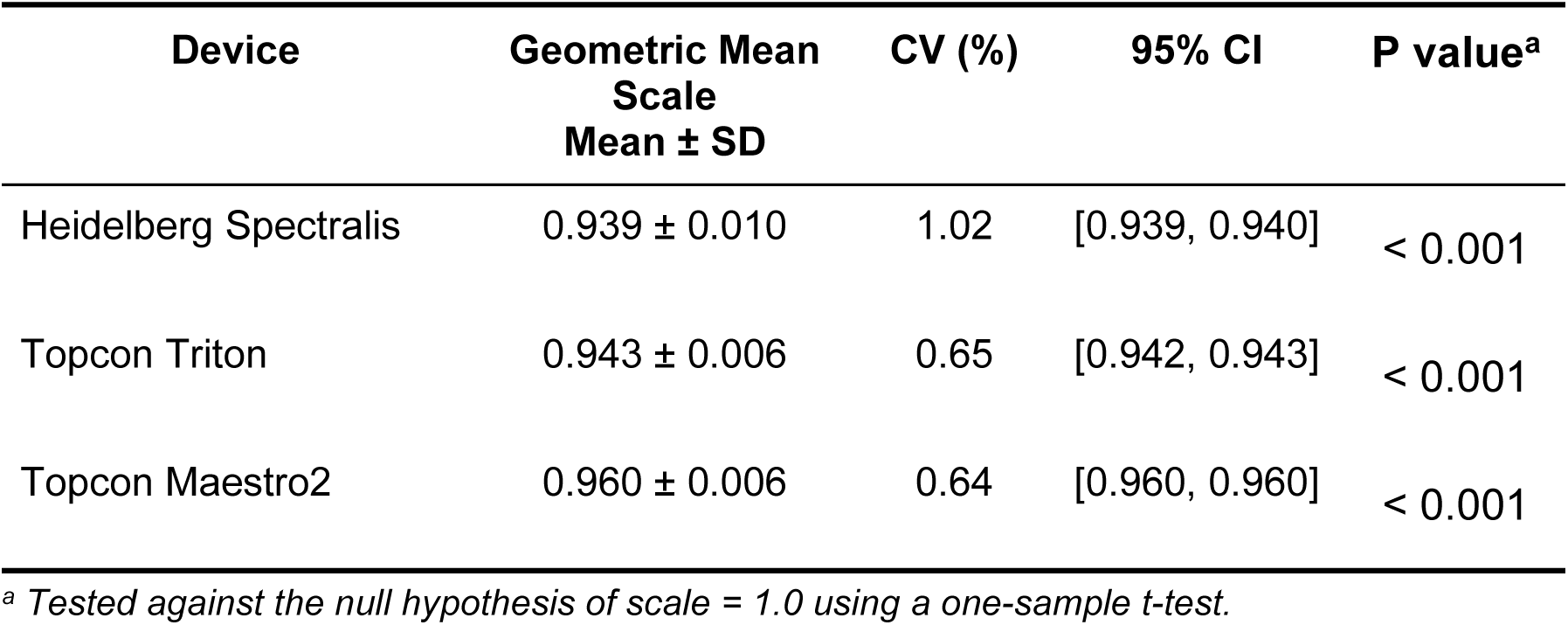
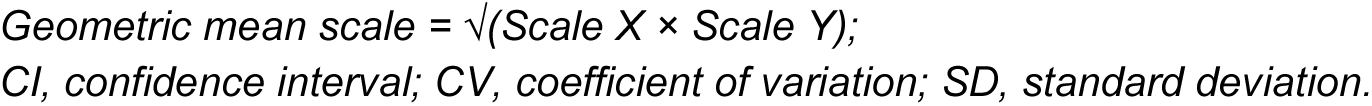
Registration-Derived Geometric Mean Scale Factors Relative to Zeiss Cirrus (n = 801 Eyes)

| Device | Geometric Mean Scale<br>Mean ± SD | CV (%) | 95% CI | P value<sup>a</sup> |
| --- | --- | --- | --- | --- |
| Heidelberg Spectralis | 0.939 ± 0.010 | 1.02 | [0.939, 0.940] | < 0.001 |
| Topcon Triton | 0.943 ± 0.006 | 0.65 | [0.942, 0.943] | < 0.001 |
| Topcon Maestro2 | 0.960 ± 0.006 | 0.64 | [0.960, 0.960] | < 0.001 |

<sup>a</sup> Tested against the null hypothesis of scale = 1.0 using a one-sample t-test.
Geometric mean scale = $\sqrt{(\text{Scale X} \times \text{Scale Y})}$ ; CI, confidence interval; CV, coefficient of variation; SD, standard deviation.

**Table 2b.**
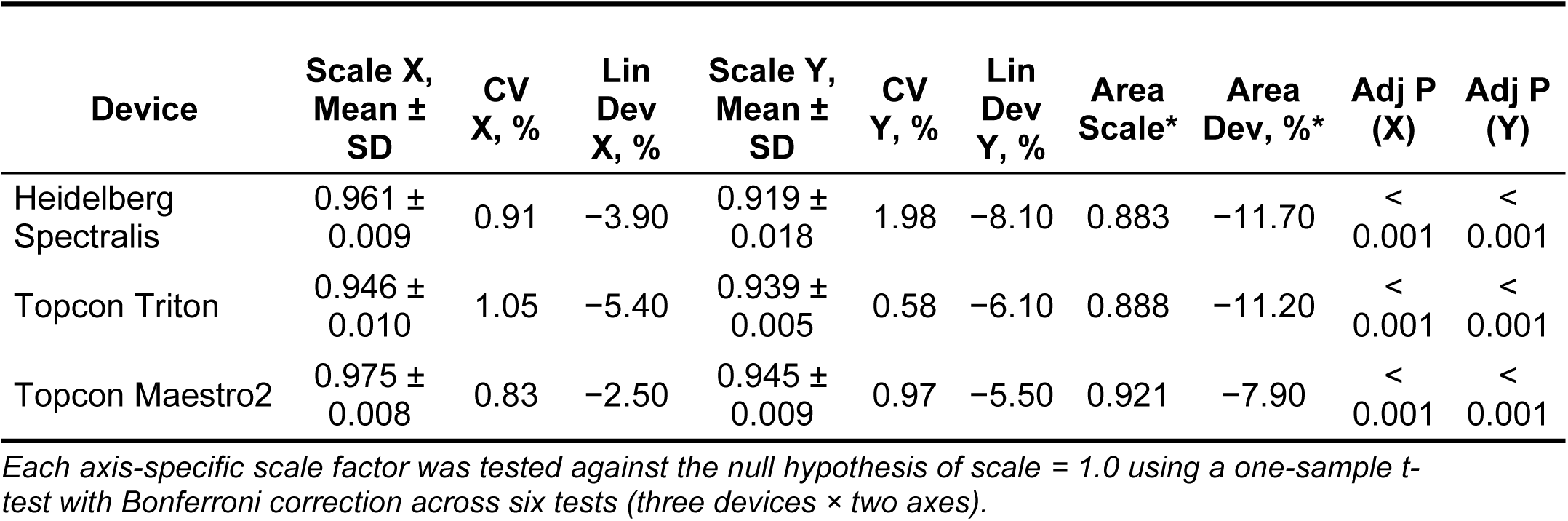
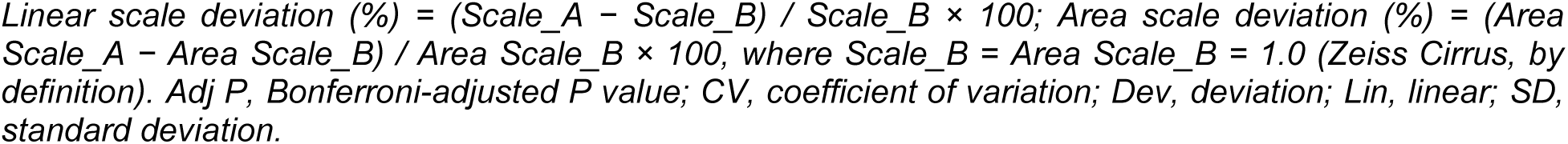
Axis-Specific Scale Factors Relative to Zeiss Cirrus (n = 801 Eyes)

| Device | Scale X,<br>Mean ±<br>SD | CV<br>X, % | Lin<br>Dev<br>X, % | Scale Y,<br>Mean ±<br>SD | CV<br>Y, % | Lin<br>Dev<br>Y, % | Area<br>Scale* | Area<br>Dev, %* | Adj P<br>(X) | Adj P<br>(Y) |
| --- | --- | --- | --- | --- | --- | --- | --- | --- | --- | --- |
| Heidelberg Spectralis | $0.961 \pm 0.009$ | 0.91 | –3.90 | $0.919 \pm 0.018$ | 1.98 | –8.10 | 0.883 | –11.70 | < 0.001 | < 0.001 |
| Topcon Triton | $0.946 \pm 0.010$ | 1.05 | –5.40 | $0.939 \pm 0.005$ | 0.58 | –6.10 | 0.888 | –11.20 | < 0.001 | < 0.001 |
| Topcon Maestro2 | $0.975 \pm 0.008$ | 0.83 | –2.50 | $0.945 \pm 0.009$ | 0.97 | –5.50 | 0.921 | –7.90 | < 0.001 | < 0.001 |

Each axis-specific scale factor was tested against the null hypothesis of scale = 1.0 using a one-sample t-test with Bonferroni correction across six tests (three devices × two axes).
*Linear scale deviation (%) = (Scale\_A - Scale\_B) / Scale\_B × 100; Area scale deviation (%) = (Area Scale\_A - Area Scale\_B) / Area Scale\_B × 100, where Scale\_B = Area Scale\_B = 1.0 (Zeiss Cirrus, by definition). Adj P, Bonferroni-adjusted P value; CV, coefficient of variation; Dev, deviation; Lin, linear; SD, standard deviation.*

Sensitivity analyses confirmed that geometric mean scale factors were not significantly associated with age or diabetes status for any device (all P > 0.01).

Axis-specific scale factors were less than 1.0 in both horizontal and vertical axes for all three devices, and the difference was statistically significant, even after Bonferroni correction, with all P values below 0.001 as reported in Table 2b.. Relative to Cirrus, Spectralis showed horizontal (X) and vertical (Y) scale factors of 0.961 ± 0.009 (−3.90% linear deviation; CV: 0.91%) and 0.919 ± 0.018 (−8.10% linear deviation; CV: 1.98%), corresponding to an area scale of 0.883 (11.70% area reduction); Triton 0.946 ± 0.010 (−5.40% linear deviation; CV: 1.05%) and 0.939 ± 0.005 (−6.10% linear deviation; CV: 0.58%), corresponding to an area scale of 0.888 (11.20% area reduction); and Maestro2 0.975 ± 0.008 (−2.50% linear deviation; CV: 0.83%) and 0.945 ± 0.009 (−5.50% linear deviation; CV: 0.97%), corresponding to an area scale of 0.921 (7.90% area reduction). The low inter-subject CVs across all axes and devices confirm that axis-specific scale factors are stable device-level properties, most likely reflecting the device-specific angular FOV.

Pairwise comparisons among the three comparison devices all reached P < 0.001 after Bonferroni correction, as shown in Table 3. Spectralis and Triton were comparable, with the smallest between-device linear deviation at-0.42% (area deviation-0.56%). Maestro2 showed larger deviations relative to both Spectralis (linear-2.19%, area-4.13%) and Triton (linear-1.77%, area-3.58%).

**Table 3.** Pairwise Comparisons of Registration-Derived Geometric Mean Scale Factors Between Comparison Devices.

| Device A | Device B | Scale (Device A) | Scale (Device B) | Linear Deviation (%) | Area Deviation (%) | P value |
| --- | --- | --- | --- | --- | --- | --- |
| Heidelberg Spectralis | Topcon Triton | 0.939 | 0.943 | -0.42 | -0.56 | < 0.001 |
| Heidelberg Spectralis | Topcon Maestro2 | 0.939 | 0.960 | -2.19 | -4.13 | < 0.001 |
| Topcon Triton | Topcon Maestro2 | 0.943 | 0.960 | -1.77 | -3.58 | < 0.001 |

*Linear deviation (%) = (Scale\_DeviceA - Scale\_DeviceB) / Scale\_DeviceB × 100.*
*Area deviation (%) = (Area Scale\_DeviceA - Area Scale\_DeviceB) / Area Scale\_DeviceB × 100, where Area Scale values are from Table 2b*
*Global comparison: one-way ANOVA, P < 0.001. Pairwise comparisons by two-sample t-test with Bonferroni correction.*

The agreement between registration-derived and phantom-measured scale factors in X and Y axes is shown in Figure 4. For all three comparison devices, registration-derived values fell within 2% of phantom-measured values in both axes. Horizontal (X) deviations were +1.59% for Spectralis, −1.77% for Triton, and −0.31% for Maestro2; vertical (Y) deviations were −0.22%, −0.53%, and −1.15%, respectively. This confirms that phantom calibration and large-scale clinical image registration yield consistent estimates of systematic inter-device spatial scale differences.

**Figure 4.**
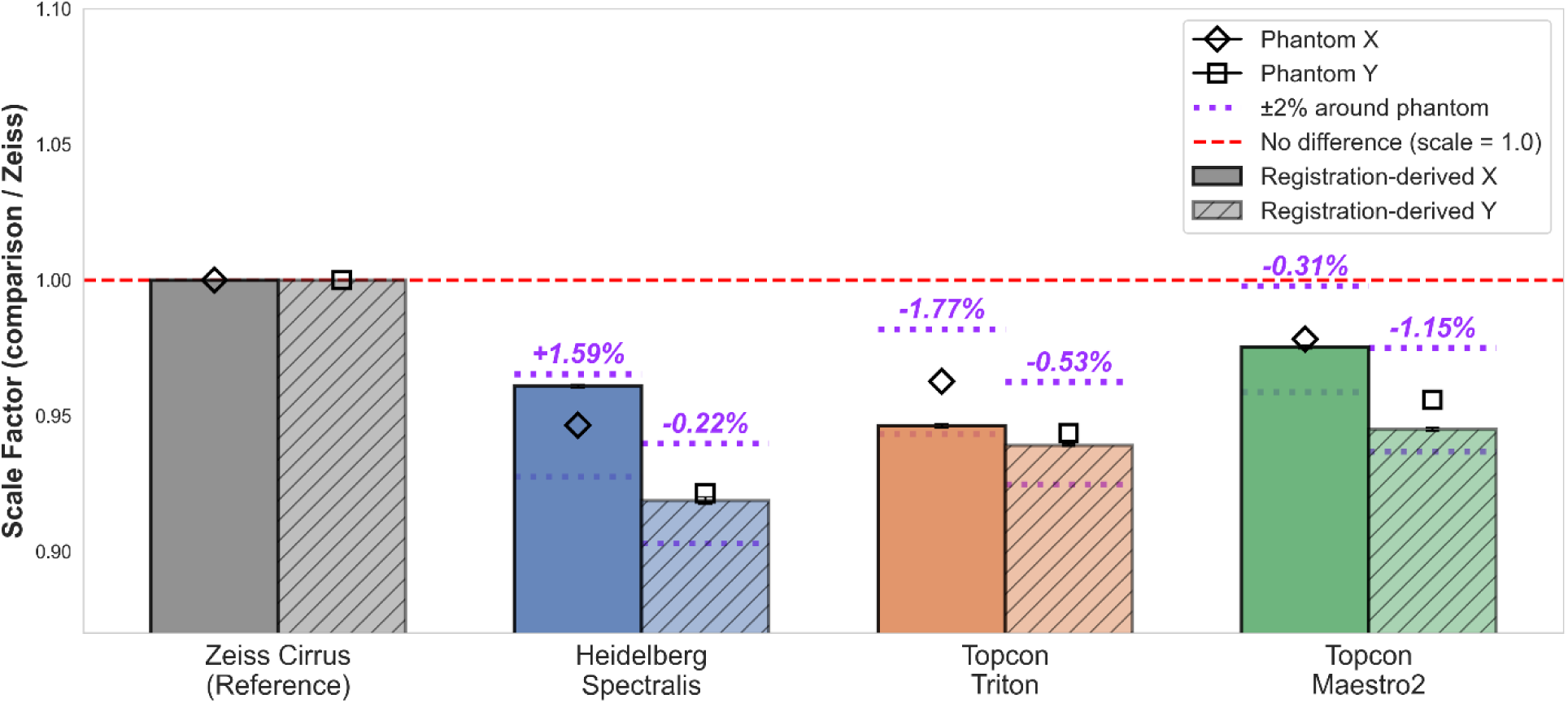
Consistency between registration-derived and phantom-measured scale factors for each device in the horizontal (X) and vertical (Y) axes. Bars show registration-derived scale factors, with solid fill for X and hatched fill for Y. Open markers show phantom-measured scale factors: diamonds for X, squares for Y. Purple dotted lines mark ± 2% around phantom-measured values; values in purple indicate the percentage difference between registration and phantom estimates. The red dashed line marks scale = 1.0.

#### 3.2.3 DICOM Metadata Analysis

DICOM-implied FOV values are shown in Figure 5. Cirrus, Triton, and Maestro2 all reported fixed DICOM pixel spacing, yielding a constant DICOM-implied FOV of 6.000 × 6.000 mm across all eyes and therefore no correlation with SE or axial length.

**Figure 5.**
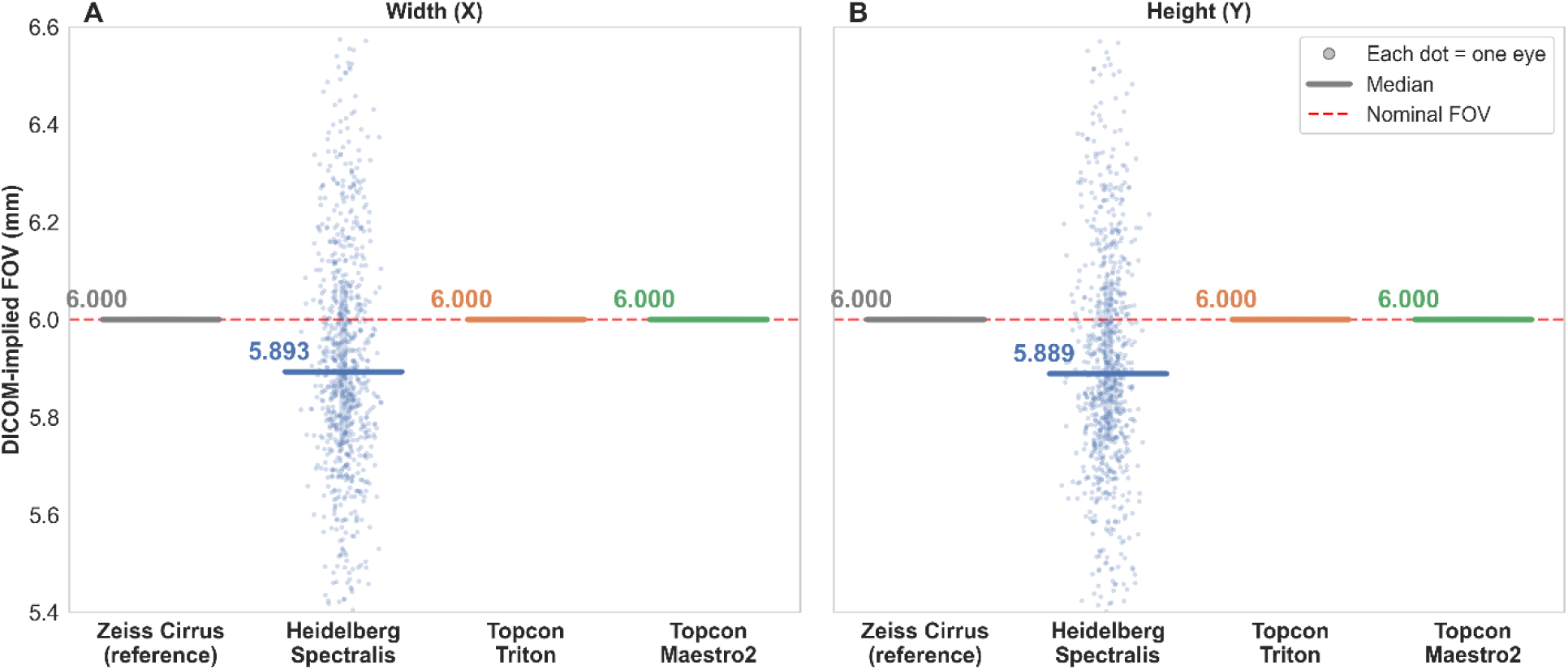
DICOM-implied field of view (FOV) per device along the (A) horizontal (X) and (B) vertical (Y) axes. Each point represents one eye, computed as the product of the stored image dimension and the corresponding pixel spacing. Horizontal lines indicate per-device medians. The red dashed line marks the nominal 6.000 mm FOV. Zeiss Cirrus, Topcon Triton, and Topcon Maestro2 reported fixed pixel spacing, yielding a constant 6.000 mm with no inter-patient variation. Heidelberg Spectralis used variable pixel spacing, producing FOV variability.

This implies no inter-device spatial scale differences; however, phantom and registration analyses both demonstrated scale differences between these devices, indicating that fixed DICOM pixel spacing does not reflect true inter-device spatial scale.

Spectralis, in contrast, employed variable pixel spacing. DICOM-implied FOV ranged from 5.212 × 5.209 to 6.743× 6.739 mm. As shown in Figure 6, this variability correlated significantly with SE, ρ = −0.635, and P < 0.001 across 274 eyes, and an equivalent correlation was found with estimated axial length. This pattern indicates that Spectralis DICOM metadata may compensate for the FOV or pixel spacing affected by the axial length.

**Figure 6.**
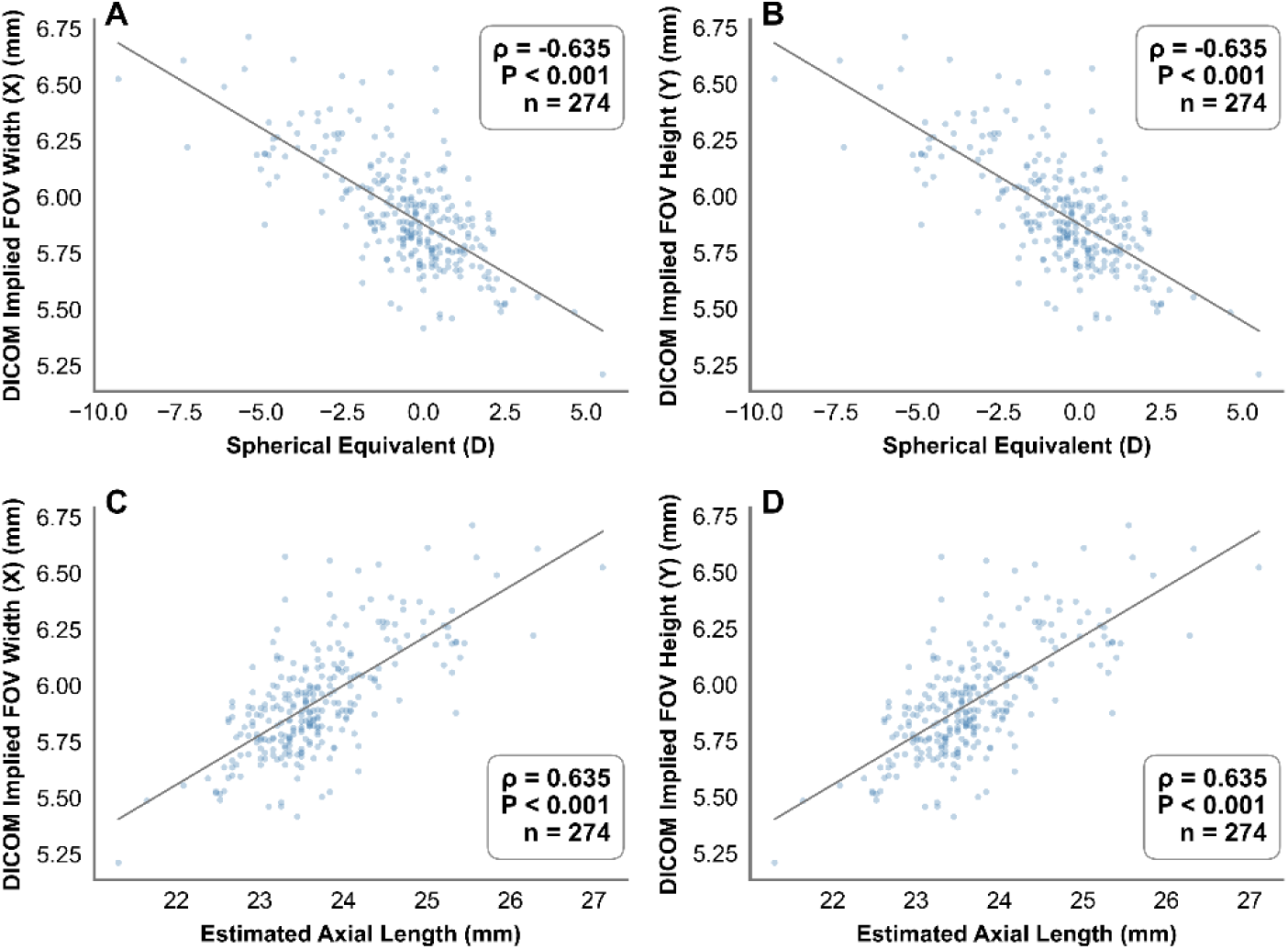
Correlation between Heidelberg Spectralis DICOM-implied field of view (FOV) and ocular biometry across 274 eyes. (A) FOV width (X) vs spherical equivalent (SE). (B) FOV height (Y) vs SE. (C) FOV width (X) vs estimated axial length. (D) FOV height (Y) vs estimated axial length. Each point is one eye. Zeiss Cirrus, Topcon Triton, and Topcon Maestro2 are not shown as their fixed DICOM pixel spacing produces identical FOV values across all eyes, precluding correlation analysis. Correlation with SE and with estimated axial length is equivalent, where estimated axial length was included for better interpretation.

## Discussion

This study provides the first systematic, multi-method characterization of FOV discrepancies among four commonly used clinical OCTA platforms under a nominally identical 6×6 mm or 20°×20° scanning protocol. Phantom calibration and *in vivo* image registration across 801 eyes converged on a consistent finding: Heidelberg Spectralis,

Topcon Triton, and Topcon Maestro2 each capture a systematically smaller FOV than Zeiss Cirrus. Registration-derived geometric mean deviations relative to Cirrus were largest for Spectralis at approximately −6.1% linear and −11.7% area deviations, intermediate for Triton at approximately −5.7% linear and −11.2% area deviations, and smallest for Maestro2 at approximately −4.0% linear and −7.9% area. These offsets were stable across eyes pooled from the three AI-READI clinical sites (UAB, UCSD, and UW), with CV ≤1.02%, indicating factory-set hardware properties rather than site-or patient-dependent variation. Sensitivity analyses using linear mixed-effects models further confirmed that scale factors were not significantly associated with age or T2DM status for any device (all p > 0.01).

Given the large sample size (N = 801 eyes) of the clinical dataset, statistical significance alone is insufficient for clinical interpretation and should be considered alongside the magnitude of the observed effects. For instance, the 0.42% linear scale difference between Spectralis and Triton, corresponding to only 0.56% in area, reached P < 0.001 yet is clinically negligible. In contrast, the area-scale deviations of each device relative to Zeiss Cirrus (7.9 – 11.7%), together with the low inter-eye variability of the scale factor (CV ≤ 2%), indicate large, consistent, device-level offsets of practical clinical consequence. Cohen’s d, a commonly used effect size measure, is similarly not informative in this context, since the inter-subject SD (≈ 0.006–0.018 primarily reflects the precision and stability of the registration-spatial scale ratio variation rather than true biological variability. Consequently, the effect size is artificially inflated by the narrow distribution of measurements rather than representing a clinically meaningful magnitude of difference.

The likely cause of these spatial scale differences lies in how each manufacturer calibrates angular FOV. Although all four devices nominally specify a 20°×20° scan angle or 6×6 mm FOV, the physical FOV depends on both the scan angle and the axial length of the reference eye assumed during internal calibration. The Littmann–Bennett formula decomposes retinal magnification into a participant-specific component determined by axial length and a device-specific component set by the system’s angular FOV calibration.^20^ Different internal reference eyes therefore produce different device-specific angular FOV under the same nominal scan setting, consistent with prior reports that manufacturers assume different internal reference axial lengths that are typically not disclosed to users.^9,26^ From phantom measurements, we roughly estimated the FOV angle from the measured linear retinal dimension (L) using the effective focal length of the eye phantom (f = 17.17 mm) according to θ = 2 arctan(L / 2f).^20^ Zeiss Cirrus yielded an angular FOV of approximately 20.0°, Spectralis 18.7°, Triton 19.0°, and Maestro2 19.3°. However, those values may slightly differ from the angular FOV at each device’s operating wavelength (≈840–1050 nm). Because the phantom lens focal length f scales as 1/(refractive index − 1), the 0.3–0.5% reduction in the refractive index of N-BK7 at the device wavelengths relative to 750 nm increases f by approximately 1.5%, which would shift these absolute angular FOV values downward by approximately 0.3°. Nevertheless, the relative spatial scale differences would remain unchanged and are independent of participant axial length.

To our knowledge, this is the first study to compare relative device-specific spatial scales across commercial OCTA platforms under a matched nominal protocol, particularly in relatively large-scale clinical validation. Prior work has addressed adjacent but distinct questions. Kraker et al. noted that manufacturers may assume different internal reference axial lengths when mapping angular FOV to millimeters, and incorporated these assumed values into magnification corrections.^26^ Their focus, however, was on correction by individual patient axial length rather than on quantifying the device-specific scale offset itself as an empirically measurable property across platforms. Ni et al. compared FOV across six ultra-widefield retinal imaging systems, with *in vivo* validation limited to a single subject.^36^ However, their focus was on system performance benchmarking rather than on quantifying the device-specific spatial scale differences that affect quantitative OCTA measurements, and each system was assessed under its own native protocol rather than under a matched nominal scan setting. This precluded direct inter-device spatial scale comparison under the identical acquisition conditions that characterize routine cross-device OCTA comparisons. The present study isolates the device-level hardware offset through an eye phantom and validates it across 801 same-eye, same-visit images acquired from 556 patients by four clinical systems. This offset cannot be corrected by patient axial length measurement alone, as long as each manufacturer’s internal calibration phantom axial length remains undisclosed.

Another related and underappreciated finding is that DICOM pixel spacing metadata may not accurately reflect the true spatial scale. Cirrus, Triton, and Maestro2 all hardcode the pixel spacings for a nominal 6.000×6.000 mm scan specification, yielding a constant implied FOV calculated from pixel spacing regardless of true retinal coverage. Consequently, any automated pipeline relying on this would systematically bias spatial-related OCTA quantitative measurements, particularly in cohorts with variable axial lengths. Instead, Spectralis uses variable pixel-spacing, with DICOM-implied FOV ranging from approximately 5.212 × 5.209 to 6.743× 6.739 mm across 801 eyes and correlating strongly with SE (and equivalently with estimated axial length). To test this variation, we performed a post hoc eye phantom experiment in Spectralis across a range of focus compensation settings from +4.01 to −8.07 D (diopters) using the device’s built-in focus compensation knob (Supplementary Table 1). The results showed that with varying focus, the DICOM pixel spacing changed from 10.77 to 13.23 μm/pixel, corresponding to an implied FOV of 5.514 to 6.774 mm, despite no change in the physical target size. Spectralis likely uses focus compensation to estimate an axial length, which is then used to adjust the pixel spacing or FOV. However, whether this compensation accurately reflects true retinal dimensions cannot be verified in our study. Despite these device-specific DICOM behaviors, the relative scale factors between Cirrus and the three comparison devices remained stable across participants and consistent with phantom findings, indicating that the device-level offsets are likely factory-set hardware properties that cannot be corrected by pixel spacing metadata alone. DICOM metadata should therefore be cautiously used before validation.

These spatial scale discrepancies across devices carry direct clinical consequences. Within-device longitudinal measurements for the same subject are largely unaffected as long as this subject’s axial length remains unchanged, but cross-device comparisons are not. A participant switching from Zeiss Cirrus to Heidelberg Spectralis could show a spurious FAZ area difference of approximately 11.7%, attributable entirely to the systems’ spatial scale difference rather than disease progression. This magnitude far exceeds the typical intra-device CV for FAZ area measurements of approximately 2% in healthy eyes.^37^ For geographic atrophy studies, where lesion enlargement rates range from 0.53 to 2.6 mm²/year ^38^, an 8–12% area bias on a typical GA lesion could obscure several months of true biological progression, a substantial confound for trials and longitudinal monitoring. Furthermore, these errors propagate directly into automated image analysis pipelines that rely on DICOM pixel spacing metadata, introducing device-dependent biases that standard domain-adaptation approaches do not address.

Based on these findings, we offer four practical recommendations regarding magnification correction for cross-device quantitative OCTA:

1. Within-subject, cross-device comparisons: When comparing the same patient across devices, the relative scale correction factors from Tables 2 and 3 are sufficient, since axial length is identical within the same eye, and there is no need to be corrected. For example, to compare a length measurement from Cirrus to Spectralis, in order to reduce the systematic device offset, Spectralis length should be multiplied by its relative linear scale factor, for example, X by 0.961 and Y by 0.919. Area measurements should be multiplied by the area scale factor, for example, 0.883.
2. Within-device, between-subject comparisons: When comparing different patients on the same device, per-patient axial length correction is sufficient since the device-level component is typically consistent.
3. Between-device, between-subject comparisons: When comparing different patients across devices, both the empirical device-level scale factors from Tables 2 and 3 and a per-patient axial length correction must be applied jointly.
4. Studies relying on DICOM pixel spacing metadata: DICOM metadata should therefore be interpreted with caution and validated before use as a spatial reference. Multi-device studies should, at a minimum, report device model and nominal scan protocol and apply the empirical scale factors reported here.

Several limitations should be acknowledged. First, absolute FOVs in millimeters depend on the phantom’s optical geometry, and the use of a Zeiss-manufactured phantom could theoretically favor Zeiss Cirrus’s absolute FOV; accordingly, these absolute values should not be interpreted as reflecting any particular patient’s absolute retinal dimension. However, the relative scale factors are directly applicable to clinical imaging and cross-device comparison. Second, only four platforms were evaluated under a single commonly used protocol.^27^ Whether similar discrepancies exist for other devices or scan sizes not included in this study warrants further investigation. Third, the correlation analyses for Spectralis used SE to represent the refractive state, with estimated axial length included for better interpretation. Because axial length was estimated from SE using a simple linear approximation, the two variables are mathematically dependent, and the correlation of DICOM-implied FOV with AL is therefore equivalent to that with SE in magnitude and does not serve as an independent validation. Nonetheless, this correlation analysis, together with the post-hoc phantom experiment, may help explain the mechanism by which Spectralis DICOM pixel spacing varies with patient refractive state. However, direct axial length measurement in future work would allow more rigorous validation. Finally, axis-specific scale factors along X and Y differed slightly across all comparison devices in both phantom and registration analyses. Although the relative scale factors were highly consistent between phantom and registration analyses, whether the observed axis differences reflect true optical anisotropy in device hardware remains uncertain. For the phantom study, this may arise from grid measurement precision, and for the registration-derived estimates, the axis differences capture the combined relative scaling between the comparison devices and the reference Cirrus and cannot be attributed to either independently. Future work with repeated measurements across multiple phantom models would help clarify whether meaningful anisotropies exist within individual devices.

In conclusion, using an eye phantom and 801 eyes from the AI-READI clinical dataset, we demonstrate that systematic FOV discrepancies exist among four commonly used clinical OCTA devices that nominally claim an identical 6×6 mm or a 20°×20° protocol. Spectralis showed the largest difference relative to Cirrus, followed by Triton and Maestro2. These device-specific scale factors were highly consistent across large-scale *in vivo* validation and agree well with the phantom study, indicating factory-set device-level hardware properties, most likely due to differences in each manufacturer’s angular-FOV. The scale factors reported here can therefore serve as practical magnification correction factors for both length-and area-based quantitative OCTA metrics to compensate for the systematic device spatial scale offset when comparing measurements across devices.

## Data Availability

All data produced in the present study are available upon reasonable request to the authors

https://aireadi.org/

## Acknowledgements

The authors acknowledge the support of Carl Zeiss Meditec for providing the eye model phantom used in this study, as well as the valuable discussions with Dr. Qinqin Zhang and Dr. Zahra Nafar (Carl Zeiss Meditec), and Dr. Mary Durbin (Topcon) throughout the project.

## Acknowledgement

## USE OF AI

The authors used Claude to assist with language editing and sentence revision. All scientific content, data analyses, and conclusions were performed and verified by the authors using original data and code.

---

**Supplementary Table 1.**
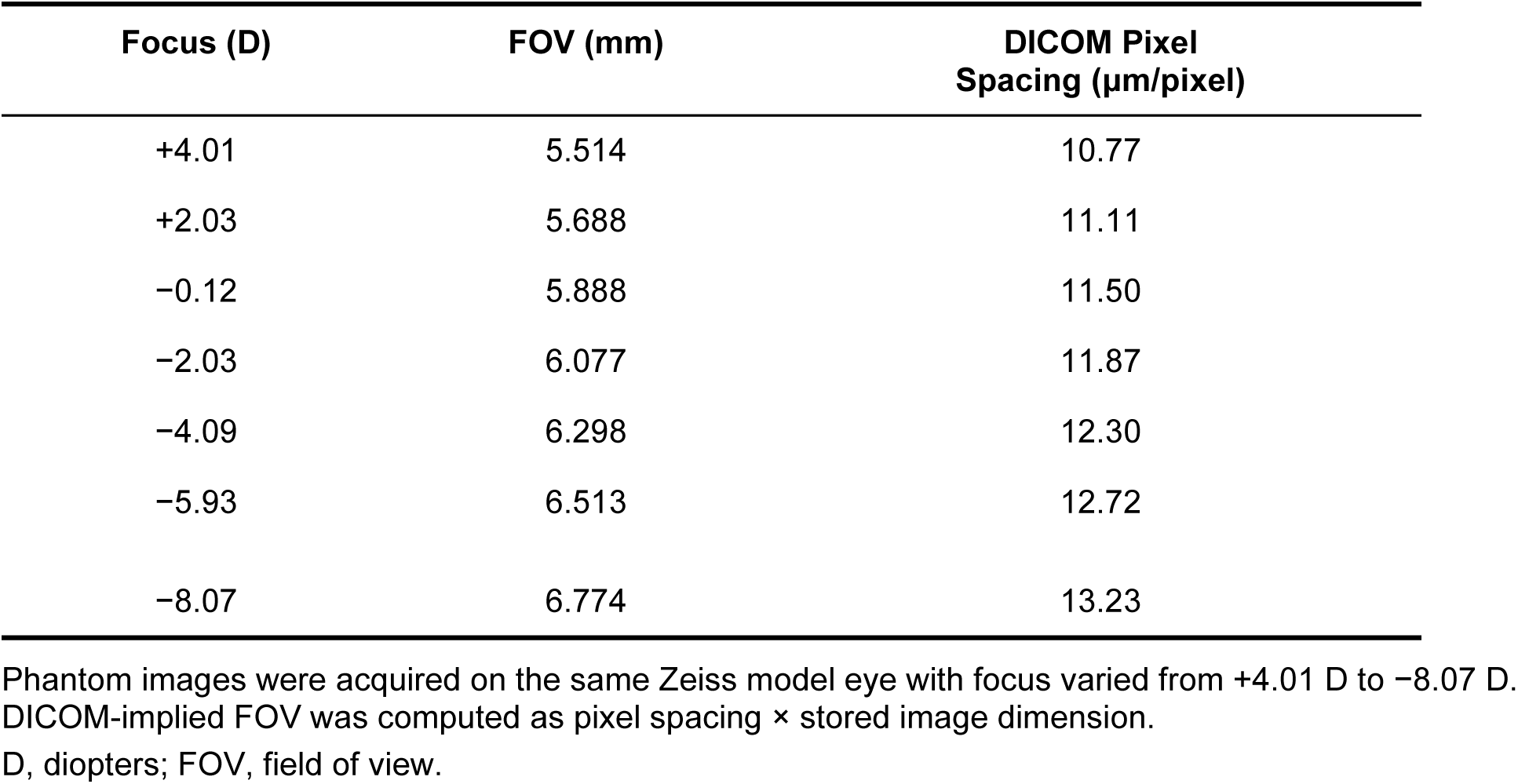
Heidelberg Spectralis DICOM-Implied Field of View and Pixel Spacing at Varying Focus Settings on the Zeiss Model Eye Phantom.

## References

1. Gong Z, Shi Y, Liu J, Zhang Y, Johnstone MA, Wang RK. Volumetric imaging of trabecular meshwork dynamic motion using 600 kHz swept source optical coherence tomography. Biomed Opt Express. 2025/01/01 2025;16(1):267-281. doi:10.1364/BOE.544521

2. Liu J, Shi Y, Gong Z, Zhang Y, Wang RK. Adaptive contour-tracking to aid wide-field swept-source optical coherence tomography imaging of large objects with uneven surface topology. Biomed Opt Express. 2024/08/01 2024;15(8):4891-4908. doi:10.1364/BOE.533399

3. Kashani AH, Chen C-L, Gahm JK, et al. Optical coherence tomography angiography: A comprehensive review of current methods and clinical applications. Progress in Retinal and Eye Research. 2017/09/01/ 2017;60:66-100. 10.1016/j.preteyeres.2017.07.002

4. de Carlo TE, Romano A, Waheed NK, Duker JS. A review of optical coherence tomography angiography (OCTA). Int J Retina Vitreous. 2015;1:5. doi:10.1186/s40942-015-0005-8

5. Tey KY, Teo K, Tan ACS, et al. Optical coherence tomography angiography in diabetic retinopathy: a review of current applications. Eye Vis (Lond). 2019;6:37. doi:10.1186/s40662-019-0160-3

6. Tsai G, Banaee T, Conti FF, Singh RP. Optical Coherence Tomography Angiography in Eyes with Retinal Vein Occlusion. J Ophthalmic Vis Res. Jul-Sep 2018;13(3):315–332. doi:10.4103/jovr.jovr_264_17

7. Shen M, Berni A, Liu J, et al. Real-World Experience With Intravitreal Pegcetacoplan for the Treatment of Geographic Atrophy in Age-Related Macular Degeneration. American Journal of Ophthalmology. 2026/01/01/ 2026;281:31-41. 10.1016/j.ajo.2025.09.006

8. Chu Z, Lin J, Gao C, et al. Quantitative assessment of the retinal microvasculature using optical coherence tomography angiography. J Biomed Opt. Jun 1 2016;21(6):66008. doi:10.1117/1.Jbo.21.6.066008

9. Sampson DM, Dubis AM, Chen FK, Zawadzki RJ, Sampson DD. Towards standardizing retinal optical coherence tomography angiography: a review. Light: Science & Applications. 2022/03/18 2022;11(1):63. doi:10.1038/s41377-022-00740-9

10. Courtie E, Kirkpatrick JRM, Taylor M, et al. Optical coherence tomography angiography analysis methods: a systematic review and meta-analysis. Scientific Reports. 2024/04/26 2024;14(1):9643. doi:10.1038/s41598-024-54306-3

11. Lujan BJ, Calhoun CT, Glassman AR, et al. Optical Coherence Tomography Angiography Quality Across Three Multicenter Clinical Studies of Diabetic Retinopathy. Transl Vis Sci Technol. Mar 1 2021;10(3):2. doi:10.1167/tvst.10.3.2

12. Cruz-Herranz A, Balk LJ, Oberwahrenbrock T, et al. The APOSTEL recommendations for reporting quantitative optical coherence tomography studies. Neurology. Jun 14 2016;86(24):2303–2309. doi:10.1212/wnl.0000000000002774

13. Hafner M, Priglinger SG, von Livonius B, Gerhardt MJ. Quantitative comparison of a novel swept-source optical coherence tomography angiography device with three established systems. Scientific Reports. 2025/06/20 2025;15(1):20129. doi:10.1038/s41598-025-04650-9

14. Arya M, Rebhun CB, Alibhai AY, et al. Parafoveal Retinal Vessel Density Assessment by Optical Coherence Tomography Angiography in Healthy Eyes. Ophthalmic Surg Lasers Imaging Retina. Oct 15 2018;49(10):S5–s17. doi:10.3928/23258160-20180814-02

15. Parrulli S, Corvi F, Cozzi M, Monteduro D, Zicarelli F, Staurenghi G. Microaneurysms visualisation using five different optical coherence tomography angiography devices compared to fluorescein angiography. Br J Ophthalmol. Apr 2021;105(4):526–530. doi:10.1136/bjophthalmol-2020-316817

16. Corvi F, Cozzi M, Barbolini E, et al. COMPARISON BETWEEN SEVERAL OPTICAL COHERENCE TOMOGRAPHY ANGIOGRAPHY DEVICES AND INDOCYANINE GREEN ANGIOGRAPHY OF CHOROIDAL NEOVASCULARIZATION. RETINA. 2020;40(5):873–880. doi:10.1097/iae.0000000000002471

17. Huang K, Su N, Tao Y, et al. Cross-Device OCTA Generation by Patch-Based 3D Multi-Scale Feature Adaption. IEEE Transactions on Emerging Topics in Computational Intelligence. 2024;8(1):641–653. doi:10.1109/TETCI.2023.3314690

18. Nouri H, Nasri R, Abtahi S-H. Addressing inter-device variations in optical coherence tomography angiography: will image-to-image translation systems help? International Journal of Retina and Vitreous. 2023/08/29 2023;9(1):51. doi:10.1186/s40942-023-00491-8

19. Untracht GR, Durkee MS, Zhao M, et al. Towards standardising retinal OCT angiography image analysis with open-source toolbox OCTAVA. Scientific Reports. 2024/03/12 2024;14(1):5979. doi:10.1038/s41598-024-53501-6

20. Bennett AG, Rudnicka AR, Edgar DF. Improvements on Littmann’s method of determining the size of retinal features by fundus photography. Graefe’s Archive for Clinical and Experimental Ophthalmology. 1994/06/01 1994;232(6):361-367. doi:10.1007/BF00175988

21. Sampson DM, Gong P, An D, et al. Axial Length Variation Impacts on Superficial Retinal Vessel Density and Foveal Avascular Zone Area Measurements Using Optical Coherence Tomography Angiography. Investigative Ophthalmology & Visual Science. 2017;58(7):3065–3072. doi:10.1167/iovs.17-21551

22. Dai Y, Zhou H, Chu Z, et al. Microvascular Changes in the Choriocapillaris of Diabetic Patients Without Retinopathy Investigated by Swept-Source OCT Angiography. Investigative Ophthalmology & Visual Science. 2020;61(3):50–50. doi:10.1167/iovs.61.3.50

23. Llanas S, Linderman RE, Chen FK, Carroll J. Assessing the Use of Incorrectly Scaled Optical Coherence Tomography Angiography Images in Peer-Reviewed Studies: A Systematic Review. JAMA Ophthalmology. 2020;138(1):86–94. doi:10.1001/jamaophthalmol.2019.4821

24. Linderman R, Salmon AE, Strampe M, Russillo M, Khan J, Carroll J. Assessing the Accuracy of Foveal Avascular Zone Measurements Using Optical Coherence Tomography Angiography: Segmentation and Scaling. Translational Vision Science & Technology. 2017;6(3):16–16. doi:10.1167/tvst.6.3.16

25. Dutt D, Yazar S, Charng J, Mackey DA, Chen FK, Sampson DM. Correcting magnification error in foveal avascular zone area measurements of optical coherence tomography angiography images with estimated axial length. Eye Vis (Lond). Aug 1 2022;9(1):29. doi:10.1186/s40662-022-00299-x

26. Kraker JA, Omoba BS, Cava JA, et al. Assessing the Influence of OCT-A Device and Scan Size on Retinal Vascular Metrics. Translational Vision Science & Technology. 2020;9(11):7–7. doi:10.1167/tvst.9.11.7

27. Gao M, Guo Y, Hormel TT, Sun J, Hwang TS, Jia Y. Reconstruction of high-resolution 6×6-mm OCT angiograms using deep learning. Biomed Opt Express. Jul 1 2020;11(7):3585–3600. doi:10.1364/boe.394301

28. Verticchio Vercellin AC, Jassim F, Poon LY-C, et al. Diagnostic Capability of Three-Dimensional Macular Parameters for Glaucoma Using Optical Coherence Tomography Volume Scans. Investigative Ophthalmology & Visual Science. 2018;59(12):4998–5010. doi:10.1167/iovs.18-23813

29. Baxter SL, de Sa VR, Ferryman K, et al. AI-READI: rethinking AI data collection, preparation and sharing in diabetes research and beyond. Nature Metabolism. 2024/12/01 2024;6(12):2210-2212. doi:10.1038/s42255-024-01165-x

30. Consortium A-R. Data from: Flagship Dataset of Type 2 Diabetes from the AI-READI Project. 2025;3.0.0. FAIRhub. 10.60775/fairhub.3

31. Kim H-S, Yu D-S, Cho HG, Moon B-Y, Kim S-Y. Comparison of predicted and measured axial length for ophthalmic lens design. PLOS ONE. 2019;14(1):e0210387. doi:10.1371/journal.pone.0210387

32. Tareen SAK, Saleem Z. A comparative analysis of SIFT, SURF, KAZE, AKAZE, ORB, and BRISK. 2018 International Conference on Computing, Mathematics and Engineering Technologies (iCoMET). 2018:1–10.

33. Alcantarilla PF, Bartoli A, Davison AJ. KAZE Features. Springer Berlin Heidelberg; 2012:214–227.

34. Cheng Y, Chu Z, Wang RK. Robust three-dimensional registration on optical coherence tomography angiography for speckle reduction and visualization. Quant Imaging Med Surg. Mar 2021;11(3):879–894. doi:10.21037/qims-20-751

35. Fischler MA, Bolles RC. Random Sample Consensus: A Paradigm for Model Fitting with Applications to Image Analysis and Automated Cartography. In: Fischler MA, Firschein O, eds. Readings in Computer Vision. San Francisco (CA): Morgan Kaufmann; 1987:726–740.

36. Ni S, Ng R, Bayhaqi Y, et al. Comparative Evaluation of Field of View Across Widefield Retinal Imaging Systems. Transl Vis Sci Technol. Nov 3 2025;14(11):20. doi:10.1167/tvst.14.11.20

37. Carpineto P, Mastropasqua R, Marchini G, Toto L, Di Nicola M, Di Antonio L. Reproducibility and repeatability of foveal avascular zone measurements in healthy subjects by optical coherence tomography angiography. British Journal of Ophthalmology. 2016;100(5):671–676. doi:10.1136/bjophthalmol-2015-307330

38. Fleckenstein M, Mitchell P, Freund KB, et al. The Progression of Geographic Atrophy Secondary to Age-Related Macular Degeneration. Ophthalmology. 2018/03/01/ 2018;125(3):369-390. 10.1016/j.ophtha.2017.08.038

